# Deep Learning-Assisted Skeletal Muscle Radiation Attenuation at C3 Predicts Survival in Head and Neck Cancer

**DOI:** 10.1101/2025.08.17.25333851

**Authors:** Felix Barajas Ordonez, Kunpeng Xie, André Ferreira, Robert Siepmann, Najiba Chargi, Sven Nebelung, Daniel Truhn, Stefaan Bergé, Philipp Bruners, Jan Egger, Frank Hölzle, Markus Wirth, Christiane Kuhl, Behrus Puladi

**Author notes:** **Corresponding author:** Kunpeng Xie, Department of Oral and Maxillofacial Surgery & Institute of Medical Informatics, University Hospital RWTH Aachen, Pauwelsstraße 30, 52074 Aachen, Germany.

## Abstract

**Background:** Head and neck cancer (HNC) patients face an increased risk of malnutrition due to lifestyle, tumor localization, and treatment effects. While skeletal muscle area (SMA) and radiation attenuation (SM-RA) at the third lumbar vertebra (L3) are established prognostic markers, L3 is not routinely available in head and neck imaging. The prognostic value of SM-RA at the third cervical vertebra (C3) remains unclear. This study assesses whether SMA and SM-RA at C3 predict locoregional control (LRC) and overall survival (OS) in HNC.

**Methods:** We analyzed 904 HNC cases with head and neck CT scans. A deep learning pipeline identified C3, and SMA/SM-RA were quantified via automated segmentation with manual verification. Cox proportional hazards models assessed associations with LRC and OS, adjusting for clinical factors.

**Results:** Median SMA and SM-RA were 36.64 cm² (IQR: 30.12–42.44) and 50.77 HU (IQR: 43.04–57.39). In multivariate analysis, lower SMA (HR 1.62, 95% CI: 1.02–2.58, p = 0.04), lower SM-RA (HR 1.89, 95% CI: 1.30–2.79, p < 0.001), and advanced T stage (HR 1.50, 95% CI: 1.06–2.12, p = 0.02) were prognostic for LRC. OS predictors included advanced T stage (HR 2.17, 95% CI: 1.64–2.87, p < 0.001), age ≥70 years (HR 1.40, 95% CI: 1.00–1.96, p = 0.05), male sex (HR 1.64, 95% CI: 1.02–2.63, p = 0.04), and lower SM-RA (HR 2.15, 95% CI: 1.56–2.96, p < 0.001).

**Conclusion:** Deep learning-assisted SM-RA assessment at C3 outperforms SMA for LRC and OS in HNC, supporting its use as a routine biomarker and L3 alternative.

## Introduction

Patients with head and neck cancer (HNC) face a heightened risk of malnutrition, due to a combination of pre-diagnosis lifestyle factors, tumor localization, and treatment-related adverse effects [1]. Prognostic assessment in HNC traditionally relies on the TNM staging system. Among additional prognostic markers, particularly p16 overexpression, has been associated with improved overall survival (OS) an disease-free survival (DFS) [2]. Head and neck imaging plays a central role in HNC management, guiding diagnosis, staging, treatment planning, and response assessment, and is routinely performed. Beyond conventional clinical and pathological prognostic markers, emerging evidence highlights body composition parameters as important biomarkers associated with worse outcomes in HNC [3,4]. However, manual assessment remains time-consuming and lacks standardization, limiting clinical adoption and driving the need for quantitative, reproducible biomarkers to improve risk stratification.

Previous studies have established that SMM depletion is associated with worse outcomes in HNC, but most relied on L3-based measurements. Grossberg et al. [3] reported that SMM depletion at L3 before and after RT was significantly associated with decreased OS (HR 1.92; 95% CI: 1.19–3.11, p = 0.007 before RT; HR 2.03; 95% CI: 1.02–4.24, p = 0.04). Nishikawa et al. [5] and Fattouh et al. [6] similarly demonstrated that sarcopenia at L3, defined by SMI, was a significant predictor of OS (HR 3.5, 95% CI: 1.2–10.0 and HR 2.1, 95% CI: 1.1–3.9, respectively). The advantage of L3-based measurements lies in the well-established cut-off values in the literature, which facilitate their use across different solid malignancies [7]. However, as L3 is only available in abdominal CT scans, these assessments are not standard in HNC, where imaging is typically confined to the head and neck region, particularly in follow-up setting.

On the other hand, C3 is almost inevitably placed in the scanning field during head and neck scans, and studies addressing its measurement have been reported. Erul et al. (2023) [8] investigated 113 patients with HNSCC undergoing radiochemotherapy (RCT) with cisplatin, evaluating sternocleidomastoid muscle volume at the C3 level. They found that pre-treatment sarcopenia was associated with lower and OS (HR 2.86; 95% CI: 1.40–5.85; p = 0.004). However, few studies have explored the prognostic value of muscle quality, rather than just quantity, as predictors of OS in patients with HNSCC.

Our study aims to assess the prognostic impact of muscle quantity, measured as skeletal muscle area (SMA), and muscle quality, assessed through skeletal muscle radiation attenuation (SM-RA), supported by a deep learning-based pipeline for automated segmentation at C3, on locoregional control (LRC) and OS in a large cohort of HNC patients undergoing multimodal treatment.

## Materials and Methods

### Study design

A total of 1355 cases were initially extracted from four public databases hosted on The Cancer Imaging Archive (TCIA): Head-Neck-PET-CT [9], TCGA-HNSC [10], CPTAC-HNSCC [11] and HEAD-NECK-RADIOMICS-HN1[12]. This retrospective study analyzed head and neck CT scans of patients with HNC from these datasets. Of the initial 1,355 cases, 904 were included in the final analysis based on the availability of clinical data and imaging covering the region of interest (C3).

Clinical variables retrieved included age, sex, tumor localization, TNM classification, therapy type, and prior surgery. Age was analyzed as a continuous variable and presented as the median with interquartile range (IQR). Tumor localization was categorized based on the affected anatomical site. For cases with CUP, a tumor stage of T0 was assigned. TNM classification followed the 7th edition of the TNM staging system. Tumor classification included T0, T1, T2, T3, and T4, with T4a and T4b grouped together. Nodal status was classified as N0, N1, and N2, with N2a, N2b, and N2c grouped as N2, and N3 as the most advanced nodal stage. Stage was categorized as Stage I, II, III, or IV, with Stage IVA and IVB grouped together. Therapy types included RT alone, surgery followed by RT, or concurrent RCT. Data on HPV status were extracted where available, but this information was unknown for 471 cases.

Outcome variables extracted included OS, measured as time to death and LRC, assessed as time to local recurrence. Among the 1355 initially identified cases, no M1 (Stage IVC) patients were present in the final cohort after applying inclusion criteria.

### Automatic Detection and Segmentation

To accurately analyze the features, we used a two-step approach to detect and segment the skeletal muscle around C3. First, we trained a detection model that uses a bounding box to localize C3 and extract its middle slices. In the middle slice, we further used a segmentation model to extract the skeletal muscle mask, including the paravertebral muscles and the sternocleidomastoid muscles bilaterally, and calculate its area in cm² (SMA) as well as the mean overall SM-RA in Hounsfield units (HU) (Figure 1).

**Figure 1.**
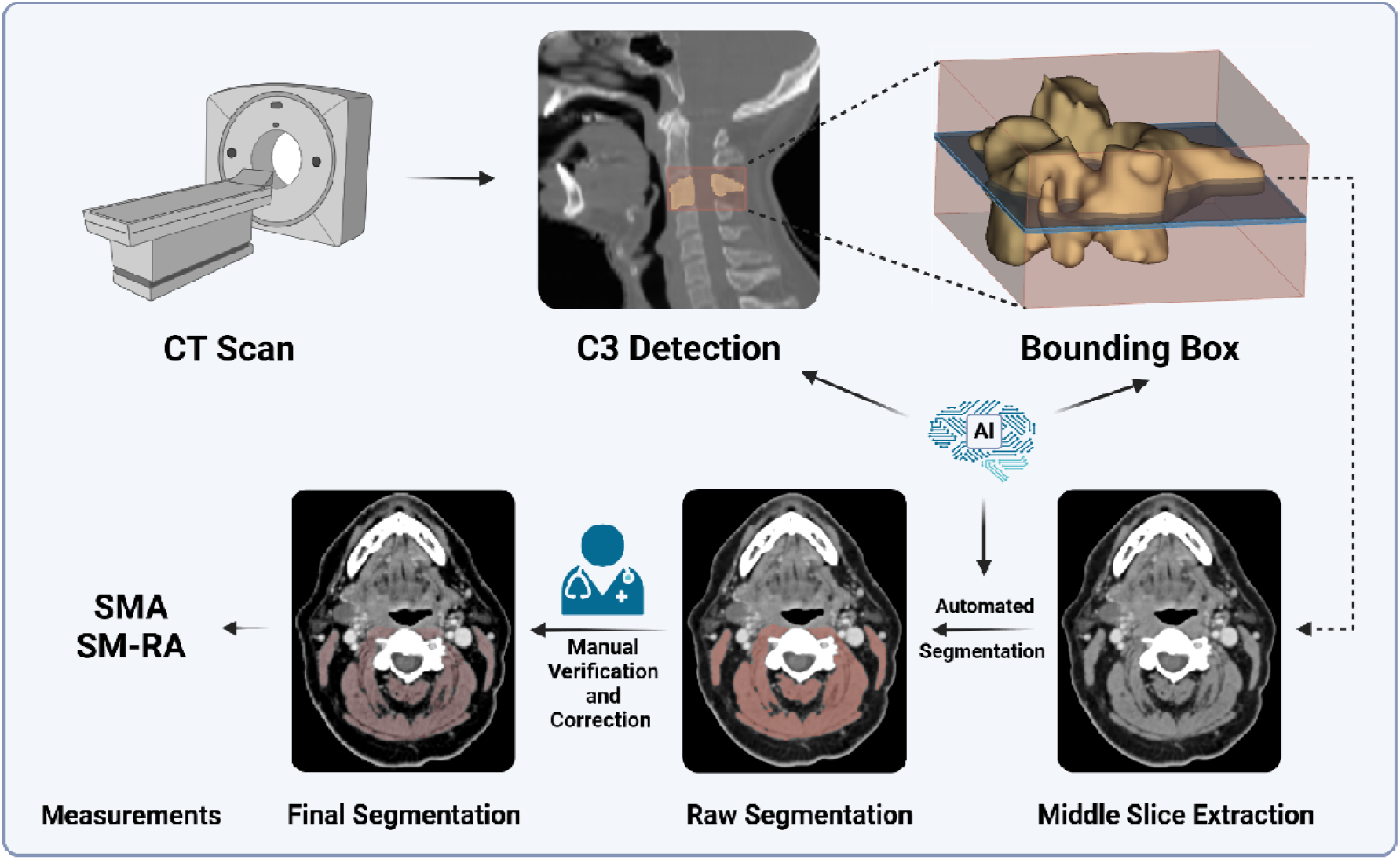
Transverse CT slice of the neck region at the level of the third cervical vertebra (C3), with the segmented skeletal muscle area shown in green, including the paravertebral muscles and the sternocleidomastoid muscles bilaterally. Created with BioRender.

### Automatic C3 Detection

We randomly selected 100 cases as a subset from 904 cases and trained a customized automatic C3 detector by manually marking the bounding boxes. Due to the high performance of the RetinaNet in the Luna16 dataset (https://luna16.grand-challenge.org/Data/), we trained this network in the MONAI framework (https://github.com/Project-MONAI/tutorials/tree/main/detection) for 3D detection. A smoothed L1 loss function and stochastic gradient descent (SGD) were used with a learning rate of 1e-2 and a weight decay of 3e-5. Training was performed using 5-fold cross-validation for 300 epochs. A visual inspection was subsequently performed by F.B.O. (a radiology resident with four years of experience in segmentation techniques) to ensure accurate segmentation of the musculature and the exclusion of other structures, such as lymph nodes. Manual adjustments were then performed by K.X. and subsequently reviewed by F.B.O. before data extraction.

### Automatic Segmentation

For the segmentation of the C3 slices selected in the previous step, we use a deep learning model pre-trained for the segmentation of muscle in C3 slices. This model is available in the repository (https://github.com/xmuyzz/C3-Segmentation). Segmentation results were reviewed by one experienced radiologists in segmentation techniques (F.B.O.). Cases with inaccuracies in C3 detection or segmentation were re-evaluated. K.X. manually corrected C3 slice selection and segmentation where necessary, and the results were reviewed and finalized by F.B.O. to ensure quality control.

This comprehensive pipeline combined automation with manual validation to optimize accuracy and reproducibility in the detection and segmentation of the C3 vertebral region.

### Statistical Analysis

The distribution of continuous variables was assessed visually using histograms and tested for normality using the Shapiro-Wilk and Kolmogorov-Smirnov tests. Variables with a non-normal distribution were presented as medians with interquartile ranges (IQR; 25th–75th percentiles), while those with approximately normal distributions were presented as means with standard deviations (SD). Kaplan-Meier survival analysis was used to estimate survival probabilities for LRC and OS, with survival curves compared across body composition groups using the log-rank test to identify statistically significant differences. Cox proportional hazards regression models were applied to evaluate LRC and OS, calculating hazard ratios (HRs) with 95% confidence intervals (CIs) and p-values. The multivariate models were adjusted for key prognostic factors in HNC, including age (≤70 vs. >70) [13], sex (male vs. female), T stage (T0–T2 vs. T3–T4), N stage (N0–N1 vs. N2–N3), therapy type (RT alone, surgery + RT, or concurrent RCT), SMA (low vs. normal/high), and SM-RA (low vs. normal/high). These adjustments allowed assessment of the independent effects of each variable on survival outcomes. Statistical significance was defined as p < 0.05, and all analyses were performed using SPSS (Version 29).

## Results

### Patient characteristics

The study cohort comprised 904 patients with a median age of 59 years (IQR: 53–66 years), predominantly men (748 patients, 82.7%). Tumor localization was primarily in the oropharynx, accounting for 738 cases (81.6%), followed by the nasopharynx (66 cases, 7.3%), larynx (34 cases, 3.8%), and hypopharynx (27 cases, 3.0%). A small subset of patients presented with CUP, which accounted for 32 cases (3.5%) and oral cavity for 7 cases (0.7%). Primary tumor size and extent, as described by the T classification, included 157 tumors (17.4%) classified as T1 and 334 tumors (36.9%) as T2. Tumors classified as T3, characterized by larger lesions or limited local extension, accounted for 237 cases (26.2%), while advanced T4 tumors were observed in 144 cases (15.9%). In terms of nodal involvement, 116 patients (12.8%) had no regional lymph node metastases (N0), while 106 patients (11.7%) presented with N1 disease. The majority of cases (635 patients, 70.2%) were classified as N2, encompassing subcategories N2a, N2b, and N2c. Advanced nodal involvement (N3) was identified in 47 patients (5.2%). Stage classification revealed a predominance of advanced disease, with 702 patients (77.6%) classified as Stage IV (including both IVA and IVB). Early stages were less common, with 11 patients (1.2%) classified as Stage I, 44 patients (4.9%) as Stage II, and 147 patients (16.3%) as Stage III.

Treatment modalities varied across the cohort. RT alone was administered to 151 patients (16.7%), and 33 patients (3.7%) received RT following surgery. Concurrent RCT was the most common approach, delivered to 607 patients (67.1%), while 113 patients (12.5%) received RCT after surgery. These data are summarized in Table 1.

**Table 1.** Patient characteristics (*n* = 904)

| Variable |  | *n* (%) |
| --- | --- | --- |
| **Age, median (IQR)** |  | 59.0 (53.0 – 59.0) |
| **Sex** | Female | 156 (17.3) |
|  | Male | 748 (82.7) |
| **Tumor Localization** | Oropharynx | 738 (81.6) |
|  | Larynx | 66 (7.3) |
|  | CUP | 32 (3.5) |
|  | Nasopharynx | 34 (3.8) |
|  | Hypopharynx | 27 (3.0) |
|  | Oral Cavity | 7 (0.8) |
| **TNM classification** |  |  |
| **T** | T0 | 32 (3.5) |
|  | T1 | 157 (17.4) |
|  | T2 | 334 (36.9) |
|  | T3 | 237 (26.2) |
|  | T4 (T4a and T4b) | 144 (15.9) |
| **N** | N0 | 116 (12.8) |
|  | N1 | 106 (11.7) |
|  | N2 (2a, 2b and 2c) | 635 (70.2) |
|  | N3 | 47 (5.2) |
| **M** | 0 | 904 (100) |
|  | 1 | 0 (0) |
| **Stage** | Stage I | 11 (1.2) |
|  | Stage II | 44 (4.9) |
|  | Stage III | 147 (16.3) |
|  | Stage IV (IVA and IVB) | 702 (77.6) |
| **HPV status** | Positive | 333 (36.8) |
|  | Negative | 100 (11.1) |
|  | Unknown | 471 (52.1) |
| **Therapy** | Radiotherapy alone | 151 (16.7) |
|  | Surgery + RT alone | 33 (3.7) |
|  | Concurrent RTC | 607 (67.1) |
|  | Surgery + RTC | 113 (12.5) |

IQR, interquartile range; CUP, Cancer of Unknown Primary, HPV (Human Papillomavirus);
RT, radiotherapy; RTC, radiochemotherapy.

### Distribution of SMA area and SM-RA

The median SMA at C3 was 36.64 cm² (IQR: 30.12–42.44 cm²), reflecting variability in muscle mass distribution across the cohort. SM-RA at C3 had a median value of 50.77 HU (IQR: 43.04–57.39 HU) (Supplementary Table 1). To assess the prognostic impact of muscle parameters, the cohort was stratified into low and normal/high SMA groups, and into low (indicative of myosteatosis) and normal/high SM-RA groups, using the 25th percentile as the cutoff for the low SMA and low SM-RA categories.

### Association between body SMA and SM-RA groups and LRC

For LRC, advanced T stage (T3-T4 vs. T0-T2) remained significantly associated with an increased hazard of locoregional failure in both univariate (HR 1.64, 95% CI 1.18–2.29, p < 0.001) and multivariate analyses (HR 1.47, 95% CI 1.04–2.09, p = 0.03). Age ≥70 years was significantly associated with reduced LRC in univariate analysis (HR 1.70, 95% CI 1.16–2.51, p = 0.001); however, this association was no longer significant in the multivariate model (HR 1.18, 95% CI 0.79–1.79, p = 0.42).

Among body composition parameters, low SMA was significantly associated with poorer LRC in univariate analysis (HR 1.83, 95% CI 1.29–2.58, p < 0.001) and remained an independent predictor in the multivariate model (HR 1.62, 95% CI 1.02–2.58, p = 0.04). Similarly, low SM-RA was strongly associated with locoregional failure in both univariate (HR 2.50, 95% CI 1.77–3.54, p < 0.001) and multivariate analyses (HR 1.89, 95% CI 1.28–2.77, p < 0.001).

While N stage (N2-N3 vs. N0-N1) and therapy type (CRT vs. RT alone) were significant in univariate analyses (p < 0.001 and p = 0.02, respectively), these factors did not retain independent significance in multivariate modeling (p = 0.06 for both) (Table 2).

**Table 2.** Association between SMA and SM-RA (as dichotomous traits) and locoregional control (*n* = 904).

| Variable | Univariate analysis |  |  | Multivariate analysis |  |  |
| --- | --- | --- | --- | --- | --- | --- |
|  | HR | 95% CI | p-value | HR | 95% CI | p-value |
| **Age**<br>*<70 vs. $\geq 70$ years* | 1.70 | [ 1.16 – 2.51] | 0.001 | 1.18 | [0.79 – 1.79 ] | 0.42 |
| **Sex**<br>*Male vs. Female* | 0.78 | [0.52 – 1.18] | 0.24 | 1.28 | [0.76 – 2.17 ] | 0.36 |
| **T Stage**<br>*$T_3$-$T_4$ vs. $T_0$-$T_2$* | 1.64 | [1.18 – 2.29] | $< \square 0.001$ | 1.47 | [1.04 – 2.09 ] | 0.03 |
| **N Stage**<br>*$N_2$-$N_3$ vs. $N_0$-$N_1$* | 0.54 | [0.39 – 0.77] | $< \square 0.001$ | 0.71 | [0.49 – 1.02 ] | 0.06 |
| **Therapy**<br>*CRT vs. RT alone* | 0.65 | [0.45 – 0.94] | 0.02 | 0.69 | [0.47 – 1.02 ] | 0.06 |
| **SMA**<br>*Low vs. Normal/High* | 1.83 | [1.29 – 2.58] | $< \square 0.001$ | 1.62 | [1.02 – 2.58 ] | 0.04 |
| **SM-RA**<br>*Low vs. Normal/High* | 2.50 | [1.77 – 3.54] | $< \square 0.001$ | 1.89 | [1.28 – 2.77] | $< \square 0.001$ |

HR, hazard ratio; CI, confidence interval; SMA, skeletal muscle area; SM-RA, skeletal muscle radiation attenuation.

### Association between body SMA and SM-RA groups and OS

For OS, advanced T stage (T3-T4 vs. T0-T2) remained a significant predictor of reduced survival in both univariate (HR 2.40, 95% CI 1.82–3.16, p < 0.001) and multivariate analyses (HR 2.15, 95% CI 1.61–2.87, p < 0.001). Age ≥70 years was significantly associated with decreased OS in univariate analysis (HR 1.94, 95% CI 1.42–2.65, p < 0.001), but its effect did not reach statistical significance in the multivariate model (HR 1.38, 95% CI 0.99–1.92, p = 0.06).

Low SM-RA was a strong predictor of worse OS, demonstrating significant associations in both univariate (HR 2.67, 95% CI 1.99–3.58, p < 0.001) and multivariate analyses (HR 2.10, 95% CI 1.52–2.91, p < 0.001). These findings were consistent with Kaplan-Meier analysis (Figure 2). In contrast, low SMA was associated with reduced OS in univariate analysis (HR 1.41, 95% CI 1.05–1.90, p = 0.02); however, this effect was not significant in the multivariate model (HR 1.39, 95% CI 0.95–2.04, p = 0.09).

**Figure 2.**
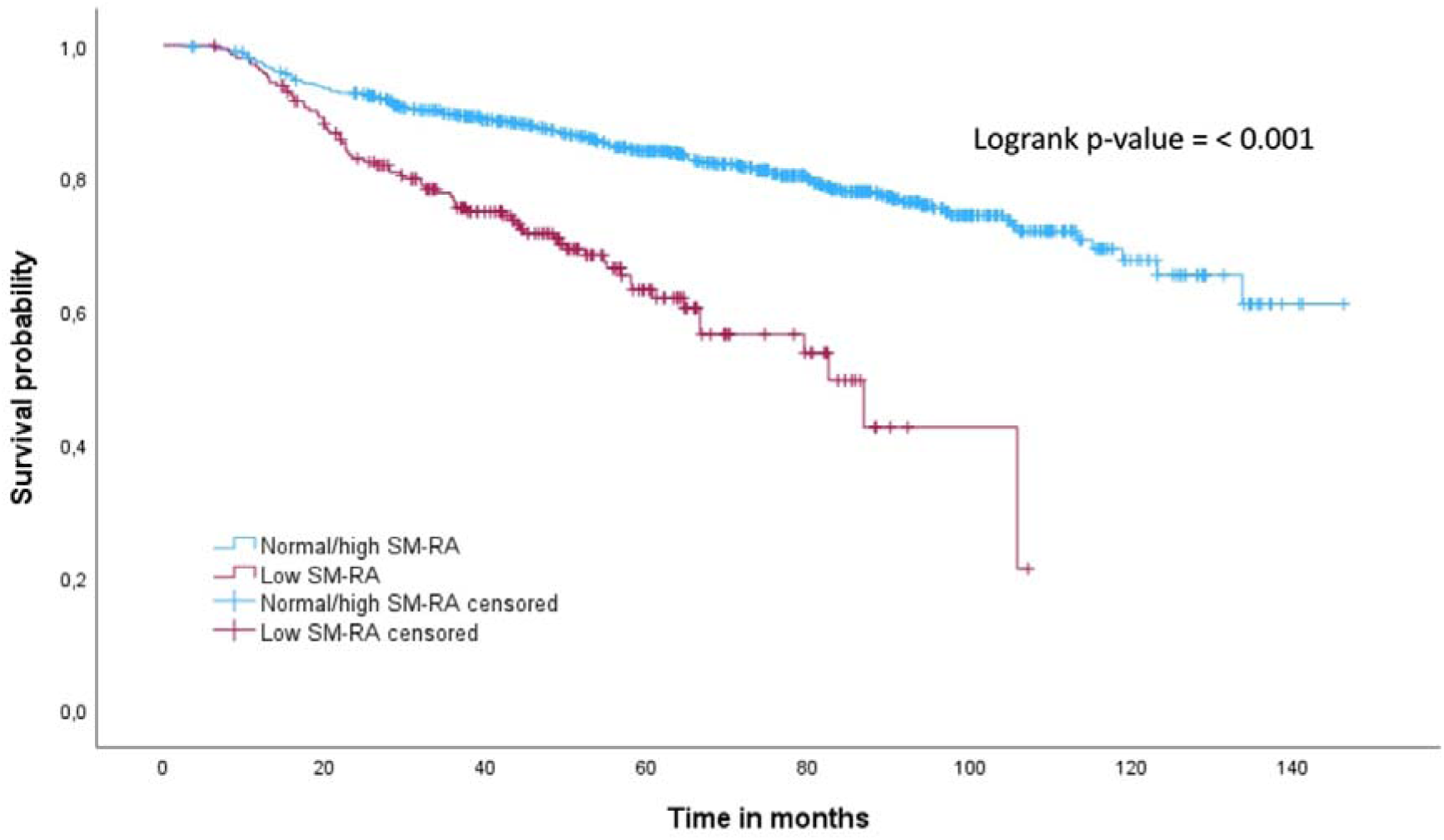
Kaplan-Meier survival analysis results for head and neck cancer (HNC) patients stratified by skeletal muscle radiation attenuation (SM-RA) into low SM-RA and normal/high SM-RA groups.

Other variables, including N stage and therapy type, did not show significant associations with OS in either univariate or multivariate analyses (p ≥ 0.07). Additionally, sex was not a significant predictor in univariate analysis but showed a significant association in multivariate modeling (HR 1.64, 95% CI 1.02–2.63, p = 0.04) (Table 3).

**Table 3.**
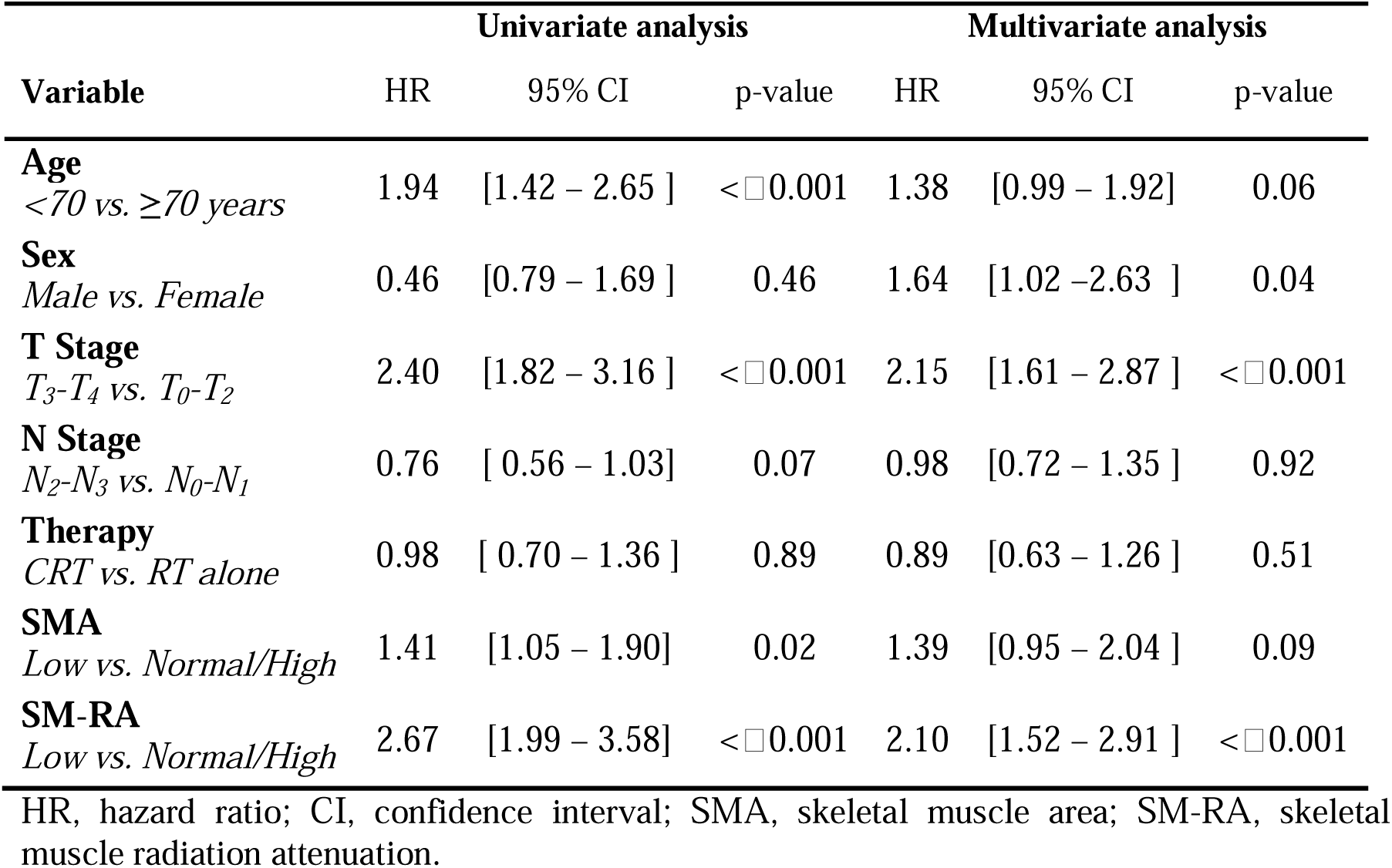
Association between SMA and SM-RA (as dichotomous traits) and overall survival (*n* = 904)

| Variable | Univariate analysis |  |  | Multivariate analysis |  |  |
| --- | --- | --- | --- | --- | --- | --- |
|  | HR | 95% CI | p-value | HR | 95% CI | p-value |
| **Age**<br><70 vs. ≥70 years | 1.94 | [1.42 – 2.65 ] | <□0.001 | 1.38 | [0.99 – 1.92] | 0.06 |
| **Sex**<br>Male vs. Female | 0.46 | [0.79 – 1.69 ] | 0.46 | 1.64 | [1.02 – 2.63 ] | 0.04 |
| **T Stage**<br>*T*<sub>3</sub>-*T*<sub>4</sub> vs. *T*<sub>0</sub>-*T*<sub>2</sub> | 2.40 | [1.82 – 3.16 ] | <□0.001 | 2.15 | [1.61 – 2.87 ] | <□0.001 |
| **N Stage**<br>*N*<sub>2</sub>-*N*<sub>3</sub> vs. *N*<sub>0</sub>-*N*<sub>1</sub> | 0.76 | [ 0.56 – 1.03] | 0.07 | 0.98 | [0.72 – 1.35 ] | 0.92 |
| **Therapy**<br>*CRT* vs. *RT alone* | 0.98 | [ 0.70 – 1.36 ] | 0.89 | 0.89 | [0.63 – 1.26 ] | 0.51 |
| **SMA**<br>Low vs. Normal/High | 1.41 | [1.05 – 1.90] | 0.02 | 1.39 | [0.95 – 2.04 ] | 0.09 |
| **SM-RA**<br>Low vs. Normal/High | 2.67 | [1.99 – 3.58] | <□0.001 | 2.10 | [1.52 – 2.91 ] | <□0.001 |

HR, hazard ratio; CI, confidence interval; SMA, skeletal muscle area; SM-RA, skeletal muscle radiation attenuation.

## Discussion

Our study evaluated both SMA and SM-RA at C3 to predict LRC and overall survival of HNC. Using a deep learning-based segmentation pipeline, we demonstrated the feasibility and reproducibility of this approach, offering an AI-driven tool for risk assessment in HNC. In multivariate analysis, lower SMA was associated with worse LRC, while lower SM-RA with decreased OS and LRC. Our findings reinforce growing evidence that muscle quality, specifically myosteatosis assessed by SM-RA, plays a critical role in oncologic outcomes [14]. In HNC, SM-RA notably outperforms SMA as a prognostic predictor, supporting its integration into routine imaging-based risk stratification. Our findings enhance clinical applicability by demonstrating how an AI-driven approach can reliably extract muscle metrics at C3 from routine head and neck imaging without additional imaging acquisition. This facilitates easier integration into clinical workflows and broader adoption of AI-driven muscle assessments in HNC.

Emerging evidence increasingly highlights the role of body composition parameters, particularly skeletal muscle mass (SMM), as key determinants of prognosis in HNC [3,4]. Grossberg et al. (2016) [3] investigated the effect of SMM depletion at both before and after radiotherapy (RT) on OS and disease control in 190 head and neck squamous cell carcinoma (HNSCC) patients. Their analysis revealed that reduced SMM at the third lumbar vertebra (L3), both prior to and following radiotherapy (RT), was independently predictive of decreased OS. Nishikawa et al. (2018) [5] further emphasized the importance of skeletal muscle index (SMI), showing that sarcopenia, as measured at the third lumbar vertebra (L3), significantly predicted OS. Consistent with these findings, Jung et al. (2019) [15] found in a prospective study involving 258 patients with advanced-stage HNSCC that sarcopenia measured before and after treatment predicted progression-free survival (PFS) and OS.

To address the limited availability of abdominal CT imaging in HNC, van Rijn-Dekker et al. [4] validated C3 as a reliable surrogate for L3 in muscle assessment. Using CSA at C3, they estimated CSA at L3 with a validated algorithm that incorporated height adjustments. Their study in 750 HNSCC patients demonstrated that sarcopenia at C3 was associated with decreased OS (HR 0.72; 95% CI: 0.56–0.93, p = 0.012) and DFS (HR 0.67; 95% CI: 0.53–0.86, p = 0.001). Endo et al. [16] analyzed 159 HNSCC patients undergoing RTC and found that low SMI at C3 was a risk factor for aspiration pneumonia, highlighting its potential clinical implications beyond survival outcomes. While anthropometric parameters were not available in the public databases used in our study, limiting calculation of CSA at L3, our semi-automated AI-driven approach provides a reproducible, fully image-based assessment independent of anthropometric data. This method enhances standardization, reduces variability, and increases clinical feasibility.

In our study, including 904 HNC cases, advanced T stage (T3-T4 vs. T0-T2) was a significant predictor of reduced LRC, retaining its significance in both univariate (HR 1.64, 95% CI 1.18–2.29, p < 0.001) and multivariate analyses (HR 1.47, 95% CI 1.04–2.09, p = 0.03). This aligns with previous literature [17], consistently highlighting tumor burden and local invasion as primary determinants of locoregional failure. Both muscle quantity and quality influenced tumor control, as low SMA remained a significant predictor of poorer LRC in multivariate analysis (HR 1.62, 95% CI: 1.02–2.58, p = 0.04). However, low SM-RA conferred an even higher risk of local tumor progression (HR 1.89, 95% CI: 1.28–2.77, p < 0.001), emphasizing muscle quality’s superior prognostic role compared to muscle mass alone. This observation is biologically plausible, as skeletal muscle serves as an energy reservoir that is mobilized during catabolic states such as cancer and chemotherapy [14]. Intramuscular adipose tissue infiltration (myosteatosis) may contribute to disease progression through the local secretion of inflammatory adipokines from adipocytes, which induces insulin resistance, impairs insulin diffusion capacity, and weakens immune defenses. This pro-inflammatory and metabolic dysregulation may create a tumor-promoting environment, ultimately facilitating neoplastic growth [14]. Given this mechanistic link, the prognostic significance of SM-RA may reflect not only muscle composition but also the systemic metabolic alterations associated with cancer-related muscle deterioration. Other factors, including therapy type, age ≥70 years, and N stage were significantly associated with LRC in univariate models but lost significance in multivariate analysis, suggesting that their effects may be mediated by other covariates.

Similarly, for OS, advanced T stage was the strongest predictor of reduced survival, in both univariate (HR 2.40, 95% CI 1.82–3.16, p < 0.001) and multivariate analysis (HR 2.15, 95% CI 1.61–2.87, p < 0.001). Low SM-RA was also associated with decreased OS, retaining its predictive value in multivariate models in multivariate models (HR 2.10, 95% CI 1.52– 2.91, p < 0.001). emphasizing muscle quality’s superior prognostic role compared to muscle mass alone.

Our results align with a meta-analysis by Aleixo et al. [14], which showed that cancer patients classified with myosteatosis had significantly lower OS compared to non-myosteatosis patients (HR 1.75, 95% CI 1.60–1.92, p < 0.00001). Importantly, the studies included in that meta-analysis primarily evaluated myosteatosis at L3, and none specifically investigated HNC. By demonstrating that SM-RA at C3 predicts OS in HNC, our study significantly extends this evidence base, underscoring the potential relevance of myosteatosis assessments beyond traditional L3-based measurements. While in our study, low SMA demonstrated an association with worse OS in univariate analysis (HR 1.41, 95% CI 1.05– 1.90, p = 0.02), its lack of significance in the multivariate model suggests muscle quality (SM-RA) may have a stronger prognostic value. Other clinical factors, such as N stage and therapy type, did not show independent associations with OS in either univariate or multivariate models, particularly advanced N stage is controversial with current literature [17]. Notably, male sex emerged as a significant predictor only after multivariate adjustment (HR 1.64, 95% CI 1.02–2.63, p = 0.04), concordant with current literature [18].

### Limitations

Our study has several limitations that should be acknowledged. First, anthropometric parameters were not available. Second, we utilized the 7th edition of the TNM classification for staging, whereas the 8th edition, which integrates p16 status for oropharyngeal carcinoma, is now the standard. However, due to the widespread lack of p16 status data in our cohort, we were unable to apply the updated staging criteria for HPV-associated cases. Given that HPV positivity is well-documented to confer a better OS, the inability to adjust for this factor represents a potential confounder that may have influenced our findings [2]. Additionally, other risk factors, such as alcohol consumption and smoking, were also not available. Future studies should incorporate HPV status to refine risk stratification further. Additionally, our analysis did not comprehensively evaluate the impact of specific systemic therapies, such as immunotherapy or monoclonal antibody therapy, which may interact with body composition parameters and influence outcomes. Furthermore, information regarding maintenance or concomitant systemic treatments was not uniformly documented, limiting our ability to assess their independent prognostic contributions. Third, the lack of established cutoff values for SMA and SM-RA at C3 remains a limitation. While C3-based measurements provide a practical alternative to L3-based body composition assessment in HNC, further validation is needed to standardize these parameters for clinical application. Nonetheless, as head and neck CT imaging is routinely performed in HNC patients, C3-based assessment remains a feasible and promising approach for integrating muscle quality and quantity into oncologic risk stratification.

### Future perspective

Future research should focus on prospective validation, establishing standardized cutoffs, and assessing its predictive value for treatment toxicity and therapy response. Beyond baseline SMA and SM-RA recent studies in other malignancies suggest that early changes in body composition parameters, particularly muscle loss during treatment, may further worsen clinical outcomes [19], reinforcing the prognostic significance of dynamic changes. However, in HNC, the prognostic impact of longitudinal SMA and particularly SM-RA assessment remains less well established. Future investigations should explore whether dynamic changes in SM-RA during treatment provide additional prognostic value beyond baseline assessments, further refining oncologic risk stratification and treatment adaptation strategies.

## Conclusion

Our findings indicate that deep learning-supported measurement of SMA and SM-RA at the C3 level is a feasible, reproducible, and clinically applicable approach for body composition assessment in HNC. Specifically, SM-RA serves as a robust biomarker and a surrogate for myosteatosis, with low SM-RA independently associated with an increased risk of locoregional recurrence and mortality. These findings reinforce their potential role in oncologic risk stratification and highlight the importance of muscle quality over muscle quantity in prognostic assessment. The integration of automated SM-RA quantification into routine imaging workflows offers a scalable, non-invasive, and time-efficient method for risk assessment, without requiring additional imaging.

## Abbreviations

C3: Third Cervical Vertebra
CI: Confidence Interval
CSA: Cross-sectional area
DFS: Disease-Free Survival
HNC: Head and Neck Cancer
HNSCC: Head and Neck Squamous Cell Carcinoma
HR: Hazard Ratio
IMAT: intermuscular adipose tissue
L3: Third Lumbar Vertebra
LRC: Locoregional control
OR: Odds Ratio
OS: Overall Survival
PFS: Progression-Free Survival
RCT: Radiochemotherapy
RT: Radiotherapy
SMA: Skeletal Muscle Area
SMI: Skeletal Muscle Index
SMM: Skeletal Muscle Mass
SM-RA: Skeletal Muscle Radiation Attenuation

## Declarations

### Author Contributions

Conceptualization, B.P. and F.B.O.; methodology, B.P.; software, K.X., A.F. and J.E.; validation, F.B.O. and K.X.; formal analysis, F.B.O., B.P., N.C., F.H., M.W. and C.K.; investigation, F.B.O.; resources, K.X. and A.F.; data curation, F.B.O., K.X. and B.P.; writing—original draft preparation, F.B.O.; writing—review and editing, B.P., F.B.O., K.X., A.F. R.S., N.C., P.B., J.E., F.H., M.W., S.N., D.T., S.B. and C.K.; visualization, R.S., K.X. and F.B.O.; supervision, B.P.; project administration, B.P.; funding acquisition, B.P.; All authors have read and agreed to the published version of the manuscript.

### Ethical Approval Statement

Ethical review and approval were not required for this study, as all analyses were conducted using publicly available, anonymized imaging datasets. This approach complies with institutional and national regulations governing the use of such data for research purposes.

### Data Availability Statement

The data presented in this study are available on reasonable request.

## Acknowledgments

The authors declare no specific acknowledgments related to this study.

## Supplementary material

**Supplementary Table 1.**
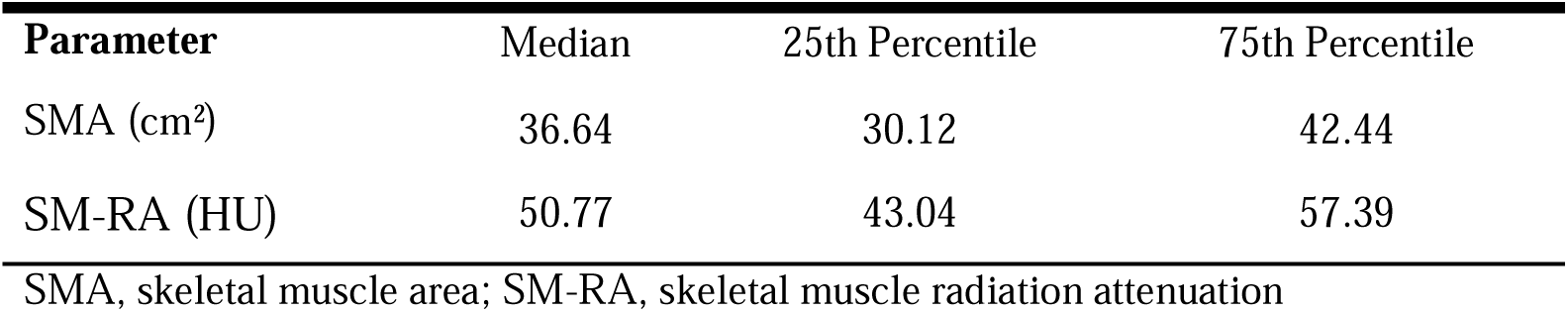
Measurements of Muscle Area and Radiodensity at Cervical Level C3 (*n* = 904).

**Suppl. Figure 1.**
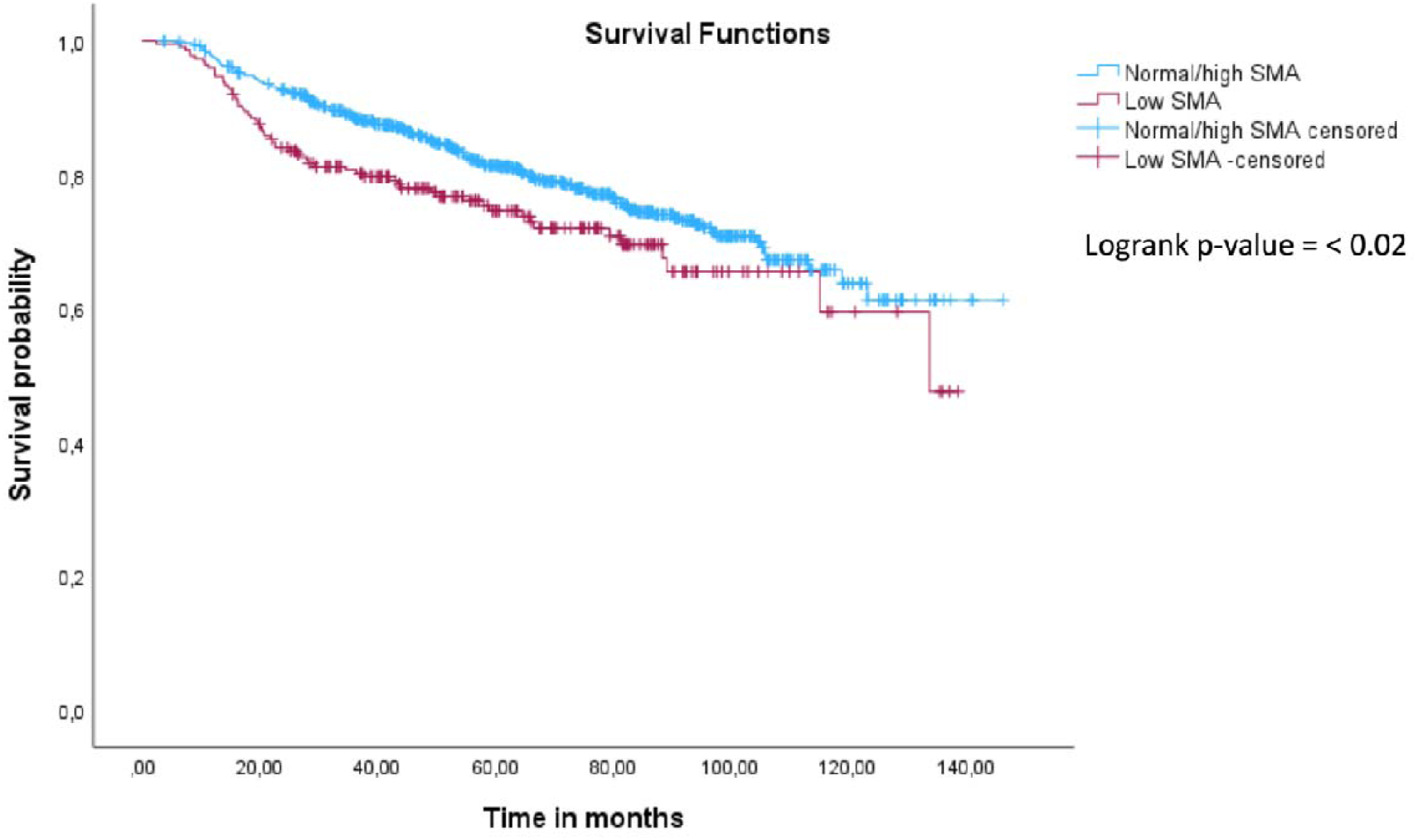
Kaplan-Meier survival analysis results for head and neck cancer (HNC) patients stratified by skeletal muscle area (SMA) into low and normal/high SMA groups.

