## Supplementary for "Deep Learning-Assisted Skeletal Muscle Radiation Attenuation at C3 Predicts Survival in Head and Neck Cancer": Supplementary material.docx

**Supplementary Table 1.**  Measurements of Muscle Area and Radiodensity at Cervical Level C3 (*n* = 904).

| **Parameter** | Median | 25th Percentile | 75th Percentile |
| --- | --- | --- | --- |
| SMA (cm²) | 36.64 | 30.12 | 42.44 |
| SM-RA (HU) | 50.77 | 43.04 | 57.39 |
| SMA, skeletal muscle area; SM-RA, skeletal muscle radiation attenuation | | | |

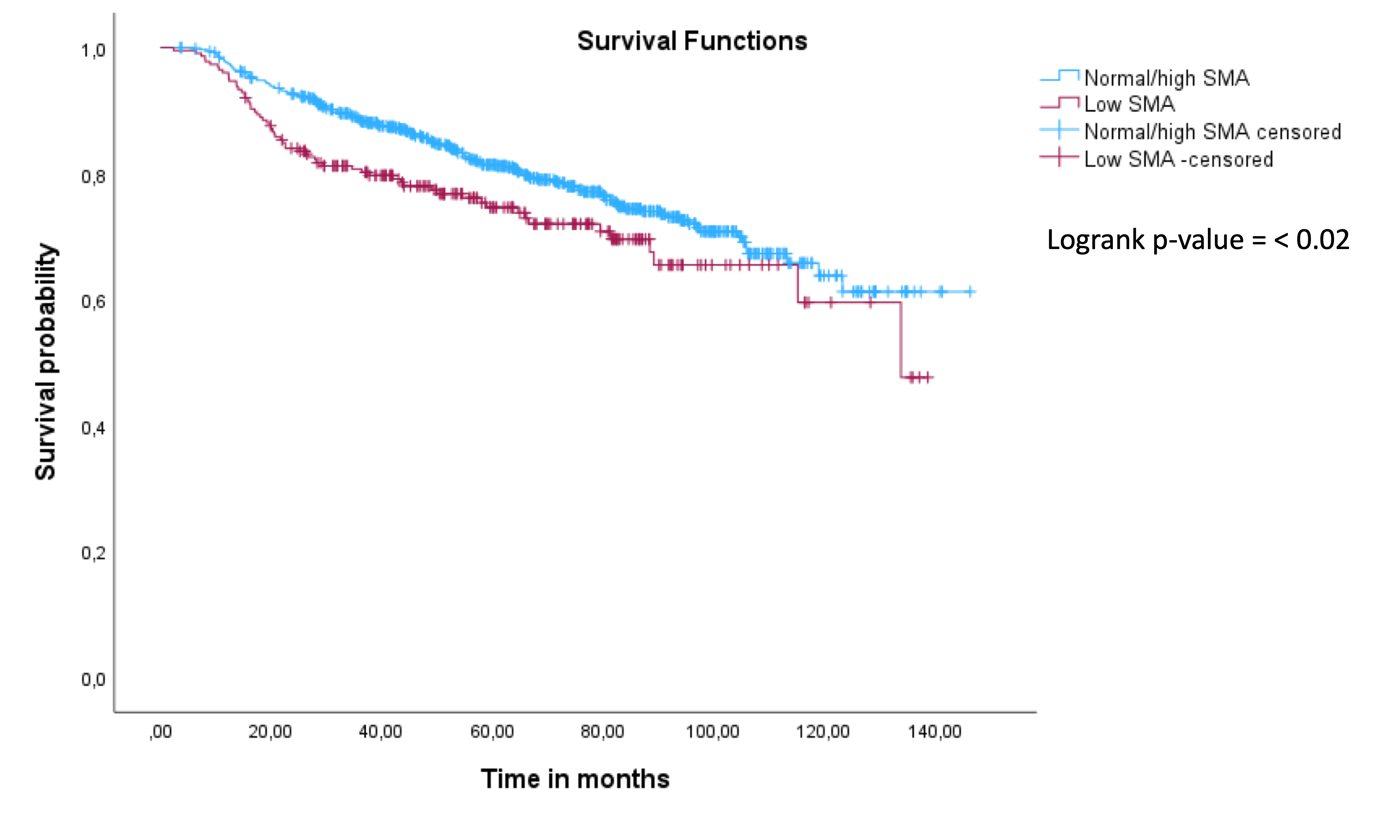

**Suppl. Figure 1.** Kaplan-Meier survival analysis results for head and neck cancer (HNC) patients stratified by skeletal muscle area (SMA) into low and normal/high SMA groups.
